# Performance of Various Lateral Flow SARS-CoV-2 Antigen Self Testing Methods in Healthcare Workers: a Multicenter Study

**DOI:** 10.1101/2022.01.28.22269783

**Authors:** V.F. Zwart, N. van der Moeren, J.J.J.M. Stohr, M.C.W. Feltkamp, R.G. Bentvelsen, B.M.W. Diederen, A.C. de Laat, E.M. Mascini, I.G.P. Schilders, H.T.M. Vlassak, H.F.L. Wertheim, J.L.A.N. Murk, J.A.J.W. Kluytmans, W. van den Bijllaardt

**Affiliations:** Microvida, Laboratory of Medical Microbiology and Immunology, location Amphia, Breda, The Netherlands; Microvida, Laboratory of Medical Microbiology and Immunology, location Elisabeth-TweeSteden, Tilburg, The Netherlands; Department of Medical Microbiology, Leiden University Medical Center, Leiden, The Netherlands; Microvida, Laboratory of Medical Microbiology and Immunology, location ZorgSaam, Terneuzen, The Netherlands; Microvida, Laboratory of Medical Microbiology and Immunology, location Bravis, Roosendaal, The Netherlands; Mijzo, Geertruidenberg, The Netherlands; Laboratory for Medical Microbiology and Immunology, Rijnstate, Arnhem, The Netherlands; Stichting Prisma, Biezenmortel, The Netherlands; GGZ Oost-Brabant, Mental Health Hospital, location Boekel, The Netherlands; Department of Medical Microbiology, Radboud University Medical Center, Nijmegen, The Netherlands; Department of medical Microbiology, University Medical Center Utrecht, Utrecht University, Utrecht, The Netherlands; Department of infection control, Amphia, Breda, The Netherlands

**Keywords:** self-testing, lateral flow SARS-CoV-2 rapid antigen test, sampling method, healthcare worker

## Abstract

**Introduction:** Rapid antigen detection tests (RDT) are suitable for large-scale testing for SARS-CoV-2 among the population and recent studies have shown that self-testing with RDT in the general population is feasible and yields acceptable sensitivities with high specificity. We aimed to determine the accuracy of two different RDT’s, with two different sample collection methods for one of the RDT’s among healthcare workers (HCW). Secondary objectives were to determine the accuracy of RDT using a viral load cut-off as proxy of infectiousness and to identify predictors for a false negative RDT.

**Methods:** Centers that participated were secondary care hospitals, academic teaching hospitals, and long-term care facilities. All HCW that met inclusion criteria were asked to perform a RDT self-test next to a regular SARS-CoV-2 nucleic acid amplification test (NAAT). Three study groups were created. Study group 1; Veritor(tm) System, Becton Dickinson, Franklin Lakes, USA (BD-RDT) with combined oropharyngeal - mid-turbinate nasal sampling, group 2; BD-RDT with mid-turbinate nasal sampling only and group 3; SD Biosensor SARS-CoV-2 Rapid Antigen Test, Roche, Basel, Switzerland (Roche-RDT) with combined oropharyngeal - mid-turbinate nasal sampling. RDT accuracy was calculated using NAAT as reference standard. For samples processed in the cobas^®^ 6800/8800 platform (Roche Diagnostics, Basel, Switzerland), established cycle threshold values (Ct-values) could be converted into viral loads. A viral load cut-off of ≥5.2 log10 SARS-CoV-2 E gene copies/ml was used as proxy of infectiousness. Logistic regression analysis was performed to identify predictors for a false negative RDT.

**Results:** In total, 7,196 HCW were included. Calculated sensitivities were 61.5% (95%CI 56.6%-66.3%), 50.3% (95%CI 42.8%-57.7%) and 74.2% (95%CI 66.4%-80.9%) for study groups 1, 2 and 3, respectively. After application of a viral load cut-off as a proxy for infectiousness for samples processed in the cobas^®^ 6800/8800 platform sensitivities increased to 82.2% (95%CI 76.6-86.9%), 61.9% (95%CI 48.8%-73.9%) and 90.2% (95%CI 76.9%-97.3%) for group 1, group 2 and group 3, respectively. Multivariable regression analysis showed that use of Roche-RDT (p <0.01), combined oropharyngeal - mid-turbinate nasal sampling (p <0.05) and the presence of COVID-19 like symptoms at the time of testing (p <0.01) significantly reduced the likeliness of a false-negative RDT result.

**Conclusion:** SARS-CoV-2 RDT has proven able to identify infectious individuals, especially when upper respiratory specimen is collected through combined oropharyngeal - mid-turbinate sampling. Reliability of self-testing with RDT among HCW seems to depend on the type of RDT, the sampling method and the presence of COVID-19 like symptoms at the time of testing.

## Introduction

Nucleic acid amplification testing (NAAT) is considered to be the reference test method for detection of SARS-CoV-2. Although less sensitive than NAAT, rapid antigen detection tests (RDT) using lateral flow assay technology are suitable for large-scale testing for SARS-CoV-2 of individuals among the general population presenting with COVID-19 like symptoms (1-5). Recent studies have shown that self-testing with RDT is feasible and yields acceptable sensitivities with high specificity (6-8). Self-testing has the potential for frequent large-scale use at a relatively low cost.

We aimed to determine the accuracy of two different RDT’s, with two different sample collection methods for one of the RDT’s. Secondary objectives were to determine the accuracy of RDT using a viral load cut-off as proxy of infectiousness and to identify predictors for a false negative RDT. Tertiary objectives were to assess the practical applicability of RDT among HCW when conducting the test themselves and the competence of HCW to interpret the RDT result themselves.

## Methods

### Study design and participants

We conducted a multicenter prospective cohort study among HCW with COVID-19 like symptoms who were analyzed for SARS-CoV-2 infection at their health care center according to local test policy. Centers that participated were secondary care hospitals, academic teaching hospitals, and long-term care facilities. The study was conducted between October 31^st^, 2020, and February 2^nd^, 2021. During this period circulation of Alpha, Delta or Omicron variants of SARS-CoV-2 was not yet observed in the Netherlands, according to national surveillance data (9).

For this study three study groups were defined. At the start of the study, all centers who were participating at that time were assigned to study group 1. Subsequent to inclusion of the required number of participants in study group 1 in accordance with the power calculation, the participating centres were switched to either study group 2 or 3. Centres were free to choose in which consecutive study group (2 or 3) they would participate. In study group 1, the BD-RDT (BD Veritor(tm) System for Rapid Detection of SARS-CoV-2, Becton Dickinson company, Franklin Lakes, USA) was used with combined oropharyngeal - mid-turbinate sampling. In study group 2, the BD-RDT was combined with mid-turbinate nasal sampling only. In study group 3, the Roche-RDT (SD Biosensor SARS-CoV-2 Rapid Antigen Test, Roche Diagnostics, Basel, Switzerland) was used with combined oropharyngeal - mid-turbinate nasal sampling.

All employees of the participating healthcare centers, both medically trained and non-medically trained, were designated as HCW. Following local test policy, all HCW with COVID-19 like symptoms made an appointment at their healthcare center of employment for a SARS-CoV-2 test based on NAAT. Every HCW aged 18 years or older and able to understand the written instructions in Dutch, was asked to participate and provide verbal informed consent.

### Sampling and self-test procedure

A sample for SARS-CoV-2 NAAT was obtained by a trained professional using the sampling method that was routinely used at the participating healthcare center. For the NAAT, nasopharyngeal or mid-turbinate sampling combined either with or without oropharyngeal sampling was assessed. Study subjects were handed a package containing the RDT device, swab stick, test medium cup, small clear plastic zip lock bag, instruction forms and question and result form. The zip lock bag was added to store the test cassette during waiting time to minimize risk of virus transmission while still allowing visual assessment of the test. Study subjects were referred to a dedicated room where they performed the self-test following the instructions. The self-test area was equipped with a mirror, table, plasticized instructions, holder for the test medium cup, chair, (chemical) waste bin, disinfection wipes, hand alcohol and sanitizer. The participants performed the tests alone, without external aid. For the instruction, question and result forms, see Supplementary Methods 1 and 2.

After conducting the self-test, a timer was to be set to 15 minutes and the short questionnaire was to be filled in. After 15 minutes the study subject was to review the test result, fill out the result on the form and take a picture of the RDT in the plastic bag, the personal barcode and the self-assessed result. The picture was to be uploaded to an anonymized and secured online cloud (Box Inc., Redwood City, USA) by scanning a QR code. The filled-out questionnaire and result form were to be deposited at the return point. A negative RDT result had no consequence on quarantine measures. In case of a positive RDT result, the same local measures applied as with a positive NAAT result.

### Nucleic Acid Amplification Tests platforms and viral load cut-off

The method of collection, transport and storage conditions of the samples were in accordance with the local protocols used in the participating centers and could vary between participating centers. Participating centers made use of their routine diagnostic laboratory protocols. Although NAAT was used as reference standard in all laboratories, the specific techniques, platforms and assays were different (Table 1). Only for real-time quantitative reverse transcription PCR (qRT-PCR) results of samples processed in the cobas^®^ 6800/8800 platform (Roche Diagnostics, Basel, Switzerland), established cycle threshold values (Ct-values) could be converted into viral loads (Supplementary Methods 3). A viral load cut-off of ≥5.2 log10 E gene copies/ml was used as proxy of infectiousness (10).

**Table 1.**

| Platforms | Molecular amplification technique | Number of tests performed (n) |
| --- | --- | --- |
| Cobas® 6800/8800 (Roche) | qRT-PCR | 3890 |
| Alinity m (Abbott) | qRT-PCR | 992 |
| Allplex™ (Seegene) | qRT-PCR | 615 |
| Panther® (Hologic) | TMA | 100 |
| BD Max™ (BD) | qRT-PCR | 63 |
| GeneXpert® (Cepheid) | qRT-PCR | 14 |
| In house / Lab developed assay | qRT-PCR | 1522 |
| Total |  | 7196 |

Overview of nucleic acid amplification test (NAAT) platforms and assays used on upper respiratory tract samples. Abbreviations: qRT-PCR, quantitative real-time reverse-transcriptase polymerase chain reaction; TMA, transcription mediated amplification.

### Sample size

We assumed the diagnostic accuracy of RDT to be lower than when performed by professionals, and based the sample size calculation on an expected sensitivity of 80%, with a margin of error of 7%, type I error of 5% and power of 90% (6). Hence, the minimum number of participants with a positive NAAT result was 125 per study group. The NAAT result positivity percentage was monitored over time and recruitment was adjusted if needed.

### Data collection and statistical analysis

Pseudonymized subject data and results of both the RDT and NAAT were collected in Castor EDC (Castor, Amsterdam, the Netherlands). Subject data consisted of sex, age, group of profession (medically trained or non-medically trained personnel), present symptoms and their answers to questions about the practicality of the test. If the RDT result and NAAT result were incongruent, the uploaded photo was assessed by a member of the research team, to acknowledge the test interpretation made by the study subject. The calculations and analyses were based on RDT results reported by the HCW.

Calculations and statistical analysis were performed using SPSS version 24 (IBM Corp. Armonk NY, USA). The overall sensitivity and specificity of RDT in the 3 study groups were calculated. When the reference NAAT was a qRT-PCR performed on the cobas^®^ 6800/8800 platform, sensitivity and specificity of the RDT could be adjusted for infectiousness. Univariable and multivariable logistic regression analyses were performed to examine whether following variables were independently associated with a false negative result in RDT as compared to NAAT: study group (categorical variable), center category (categorical variable), sex (categorical variable), age (continuous variable), group of profession (categorical variable) and current COVID-19 related symptoms (categorical variable). Variables were included in the multivariable analysis when p <0,2 in the univariable analysis. Invalid RDT results were excluded when determining sensitivity and specificity of self-testing and interpreted as not false negative when determining the variables associated with a false negative RDT result.

### Ethics

The planning, conduction, documentation and reporting of the study were in line with the Declaration of Helsinki, as revised in 2013. The study protocol was reviewed by the Dutch ‘Medical research Ethics Committees United’ (MEC-U) and was judged to be beyond the scope of the Dutch medical scientific research act (WMO) (MEC-U subject: W20.250). A waiver of written informed consent was granted as handling of documents obtained from possibly infectious participants was considered a potential safety hazard.

## Results

### Study design and baseline characteristics

During the study period, 7,495 HCW were considered eligible for participation of whom 7,196 HCW were included (Fig. 1). Participants were excluded if there was a missing RDT result (3%), NAAT result (<1%), study ID (<1%) or date of testing (<1%). In study group 1, BD-RDT with combined oropharyngeal - mid-turbinate nasal sampling, 3,255 HCW were included. In study group 2, BD-RDT with mid-turbinate nasal sampling only, 1,729 HCW were included. In study group 3, Roche-RDT with combined oropharyngeal - mid-turbinate nasal sampling, 2,212 HCW were included. Baseline characteristics of the three study groups were comparable between all groups and are presented in Table 2.

**Table 2.**

|  | Study group |  |  | Overall |
| --- | --- | --- | --- | --- |
|  | Group 1* | Group 2** | Group 3*** |  |
| Age in years, median (IQR) | 40 (30-52) | 39 (29-51) | 40 (30-52) | 40 (30-52) |
| Gender, n (%) |  |  |  |  |
| Female | 2759 (85,9) | 1377 (80,3) | 1820 (83,1) | 5956 (83,7) |
| Male | 452 (14,1) | 338 (19,7) | 369 (16,9) | 1159 (16,3) |
| Group of profession, n (%) |  |  |  |  |
| Medical | 2435 (76,9) | 1189 (71,5) | 1716 (78,5) | 5340 (76,1) |
| Non-medical | 731 (23,1) | 474 (28,5) | 470 (21,5) | 1675 (23,9) |
| Supportive | 422 (13,3) | 289 (17,4) | 282 (12,9) | 993 (14,2) |
| Office | 290 (9,2) | 177 (10,6) | 181 (8,3) | 648 (9,2) |
| Volunteer | 19 (0,6) | 8 (0,5) | 7 (0,3) | 34 (0,5) |
| Currently symptoms of COVID-19, n (%) |  |  |  |  |
| Yes | 2697 (84,3) | 1277 (74,8) | 1693 (77,1) | 5667 (79,8) |
| No | 503 (15,7) | 431 (25,2) | 502 (22,9) | 1436 (20,2) |

\*BD-RDT with combined oropharyngeal - mid-turbinate nasal sampling. \*\* BD-RDT with mid-turbinate nasal sampling only. \*\*\*Roche-RDT with combined oropharyngeal - mid-turbinate nasal sampling.
Abbreviations: IQR, interquartile range; COVID-19, coronavirus disease 2019.

**Fig. 1.**
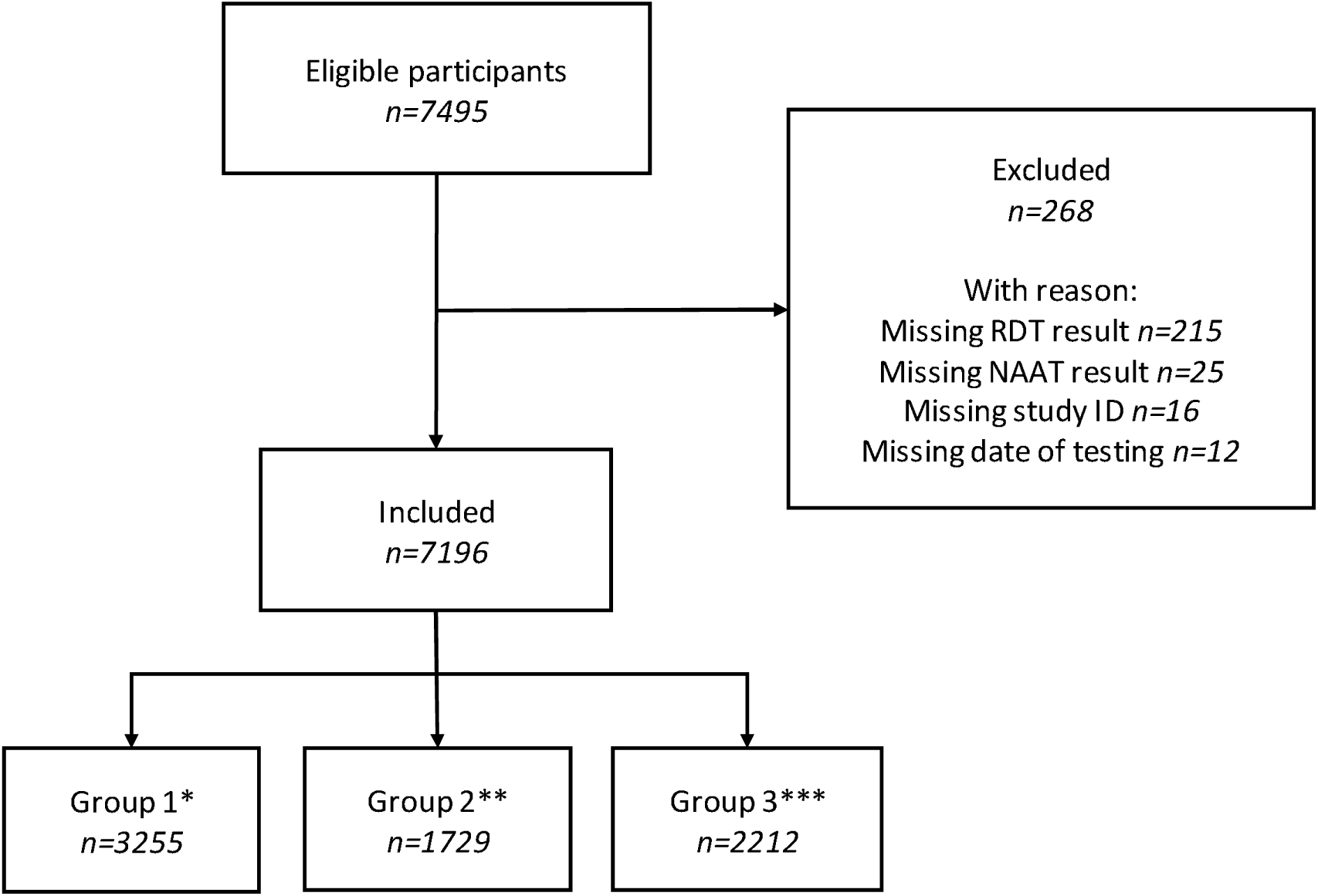
*BD-RDT with combined oropharyngeal - mid-turbinate nasal sampling. ** BD-RDT with mid-turbinate nasal sampling only. ***Roche-RDT with combined oropharyngeal - mid-turbinate nasal sampling. Abbrevations: RDT, rapid antigen detection test; NAAT, nucleic acid amplification test.

An overview of the participating centers, corresponding number of inclusions in the various study groups are presented in Table 3. All but two centers started in study group 1 and then decided to proceed to study group 2 or 3 or to stop participation. One center started in study group 2, one center in study group 3 and one center decided to stop further participation after the required numbers in study group 1 had been reached.

**Table 3.**

|  | Total inclusions<br>(n) | Inclusions<br>group 1* (n) | Inclusions<br>group 2** (n) | Inclusions<br>group 3*** (n) |
| --- | --- | --- | --- | --- |
| Secondary care hospitals |  |  |  |  |
| Amphia ziekenhuis | 1814 | 812 | 0 | 1002 |
| Bravis ziekenhuis | 1199 | 681 | 518 | 0 |
| ZorgSaam ziekenhuis | 343 | 343 | 0 | 0 |
| Admiraal de Ruyter ziekenhuis (ADRZ) | 167 | 129 | 38 | 0 |
| Canisius Wilhelmina ziekenhuis (CWZ) | 847 | 386 | 0 | 461 |
| Rijnstate ziekenhuis | 357 | 0 | 0 | 357 |
| Academic teaching hospitals |  |  |  |  |
| Radboud Universitair Medisch Centrum (RUMC) | 340 | 47 | 293 | 0 |
| Leids Universitair Medisch Centrum (LUMC) | 277 | 0 | 277 | 0 |
| Long-term care facilities |  |  |  |  |
| MIJZO | 485 | 289 | 196 | 0 |
| Thebe | 342 | 215 | 0 | 127 |
| PRISMA | 553 | 173 | 380 | 0 |
| GGZ Oost-Brabant | 414 | 149 | 0 | 265 |
| Ons Tweede Thuis | 58 | 31 | 27 | 0 |
| Total | 7196 | 3255 | 1729 | 2212 |

\*BD-RDT with combined oropharyngeal - mid-turbinate nasal sampling. \*\* BD-RDT with mid-turbinate nasal sampling only. \*\*\*Roche-RDT with combined oropharyngeal - mid-turbinate nasal sampling.

### RDT sensitivity and specificity

In study group 1, 411 (12.6%) participants had a NAAT confirmed SARS-CoV-2 infection and 253 (7.8%) participants had a positive result in the RDT. RDT sensitivity and specificity were 61.5% (95%CI 56.6% to 66.3%) and 99.9% (95%CI 99.6% to 100%), respectively (Table 4). In study group 2, 191 (11.0%) participants had a NAAT confirmed SARS-CoV-2 infection and 96 (5.6%) participants had a positive result in the RDT resulting in a sensitivity and specificity of 50.3% (95%CI 42.8% to 57.7%) and 99.7% (95%CI 99.3% to 99.9%), respectively. In study group 3, 152 (6.9%) participants had a NAAT confirmed SARS-CoV-2 infection and 119 (5.4%) participants had a positive result in the RDT resulting in a sensitivity of 74.2% (95%CI 66.4% to 80.9%) and specificity of 99.7% (95%CI 99.3% to 99.9%).

**Table 4.**

|  | Study group |  |  | Overall |
| --- | --- | --- | --- | --- |
|  | Group 1* | Group 2** | Group 3*** |  |
| NAAT result, n (%) |  |  |  |  |
| Positive | 411 (12,6) | 191 (11,0) | 152 (6,9) | 754 (10,5) |
| Negative | 2844 (87,4) | 1538 (89,0) | 2060 (93,1) | 6442 (89,5) |
| RDT result |  |  |  |  |
| Positive | 253 (7,8) | 96 (5,6) | 119 (5,4) | 468 (6,5) |
| Negative | 2965 (91,1) | 1591 (92,0) | 2073 (93,7) | 6629 (92,1) |
| Invalid | 37 (1,1) | 42 (2,4) | 20 (0,9) | 99 (1,4) |
| Sensitivity RDT overall, % (95% CI) | 61,5 (56,6-66,3) | 50,3 (42,8-57,7) | 74,2 (66,4-80,9) | 61,3 (57,7-64,8) |
| Specificity RDT overall, % (95% CI) | 99,9 (99,6-100) | 99,7 (99,3-99,9) | 99,7 (99,3-99,9) | 99,8 (99,6-99,9) |
| *BD-RDT with combined oropharyngeal - mid-turbinate nasal sampling. ** BD-RDT with mid-turbinate nasal sampling only. ***Roche-RDT with combined oropharyngeal - mid-turbinate nasal sampling. Abbreviations: NAAT, nucleic acid amplification test; RDT, rapid antigen detection test; CI, confidence interval. |  |  |  |  |

For qRT-PCR results of samples processed in the cobas^®^ 6800/8800 platform, a viral load cut-off was used as a proxy for infectiousness. In study group 1, 304 (13.8%) of all 2,210 samples processed on the cobas^®^ 6800/8800 platform had a positive qRT-PCR result and 233 (76.6%) of these, had a viral load above the cut-off. This resulted, after application of a viral load cut-off as a proxy for infectiousness, in a sensitivity and specificity of 82.2% (95%CI 76.6 to 86.9%) and 99.3% (95%CI 98.8% to 99.6%) for the BD-RDT with combined oropharyngeal - mid-turbinate nasal sampling, respectively (Table 5). The same procedure was applied for study group 2 and 3. In study group 2, 86 (12.6%) of all 1,992 samples run on the cobas^®^ 6800/8800 platform had a positive qRT-PCR result and of those, 65 (75.6%) had a viral load above the cut-off. After application of a viral load cut-off as a proxy for infectiousness, this resulted in a sensitivity and specificity of 61.9% (95%CI 48.8% to 73.9%) and 99.8% (95%CI 99.1% to 100%) for the BD- RDT with mid-turbinate nasal sampling only, respectively. In study group 3, 56 (5.6%) of all 997 samples run on the cobas^®^ 6800/8800 platform had a positive qRT-PCR result and of those, 41 (73.2%) had a viral load above the cut-off. This resulted, after application of a viral load cut-off as a proxy for infectiousness, in a sensitivity and specificity of 90.2% (95%CI 76.9% to 97.3%) and 99,4% (95%CI 99.1% to 99.6%) for the Roche-RDT with combined oropharyngeal - mid-turbinate nasal sampling, respectively.

**Table 5.**
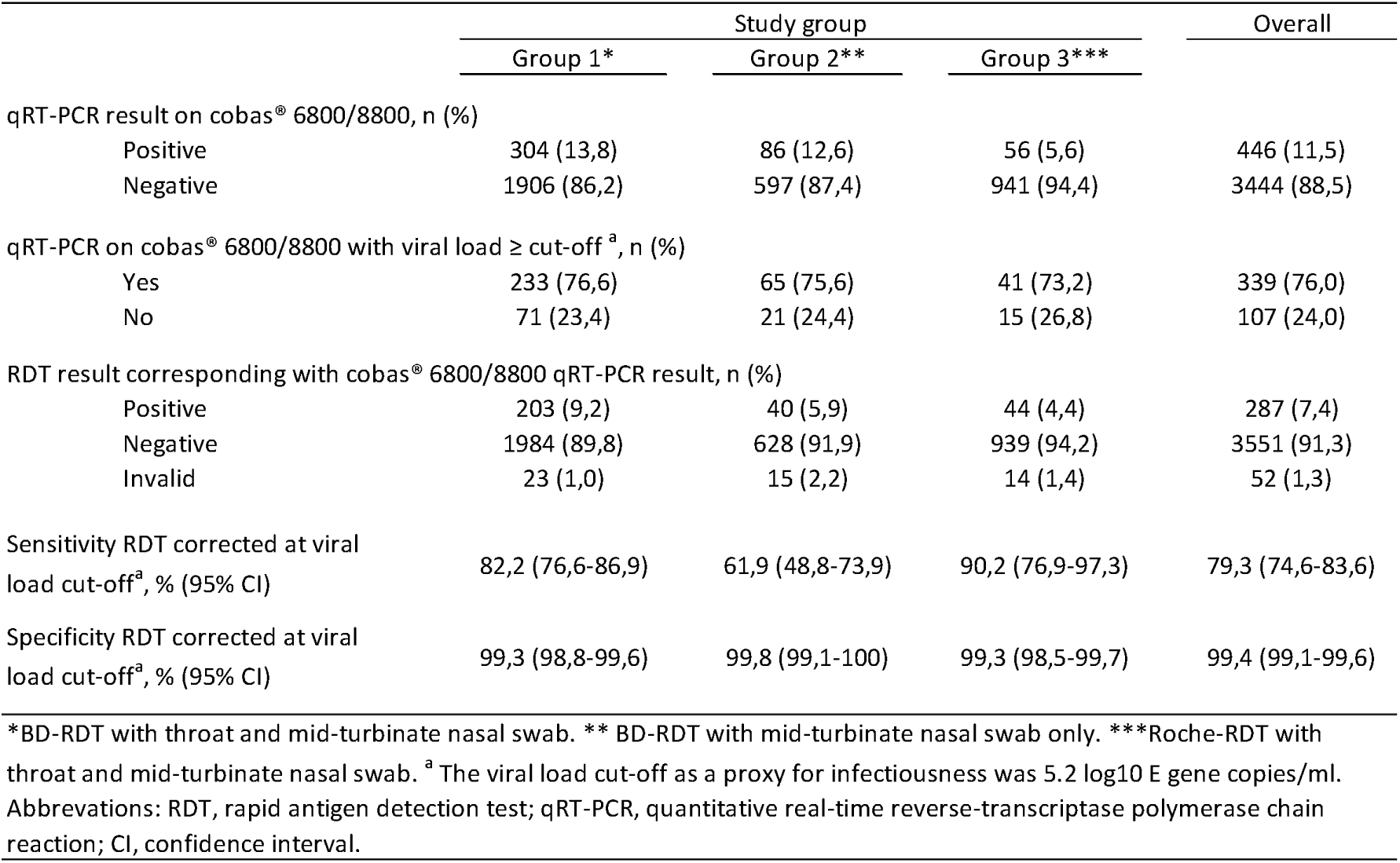

### Multivariable analysis

The relation between a false-negative RDT (negative RDT result in case of a positive NAAT, irrespective of viral load) was assessed for seven variables by conducting a univariable logistic regression analysis (Table 6). Out of these seven variables, three were associated with the occurrence of a false-negative RDT result with a p-value <0.2 and were included in the multivariable analysis (Table 6). The type of RDT was independently associated with the occurrence of a false-negative RDT result (p <0,01). Participants that assessed a Roche-RDT were significantly less likely to have a false-negative RDT result compared to participants that assessed a BD-RDT. Also, the method of sampling was independently associated with the occurrence of a false-negative RDT result (p <0,05). Participants that performed combined oropharyngeal – mid-turbinate nasal sampling were significantly less likely to have a false-negative RDT result compared to participants that performed mid-turbinate nasal sampling only. As the third variable, the presence of COVID-19 like symptoms at the time of testing was independently associated with the non-occurrence of a false-negative RDT result (p <0.01).

**Table 6.**

|  | Univariable analysis |  | Multivariable analysis |  |
| --- | --- | --- | --- | --- |
|  | Odds ratio (95% CI) | p-value | Odds ratio (95% CI) | p-value |
| Roche-RDT<sup>a</sup> | 0,496 (0,333-0,739) | 0,001 | 0,538 (0,351-0,824) | 0,004 |
| Combined oropharyngeal and mid-turbinate nasal sampling method<sup>a</sup> | 0,582 (0,417-0,812) | 0,001 | 0,697 (0,489-0,995) | 0,047 |
| Center category |  |  |  |  |
| Academic teaching hospital |  | 0,485 |  |  |
| Secondary care hospital | 0,733 (0,398-1,351) | 0,320 |  |  |
| Long-term care facility | 0,680 (0,362-1,277) | 0,231 |  |  |
| Female<sup>a</sup> | 1,152 (0,749-1,771) | 0,519 |  |  |
| Age (years) | 0,997 (0,986-1,008) | 0,604 |  |  |
| Medically trained personnel<sup>a</sup> | 0,799 (0,554-1,151) | 0,228 |  |  |
| Symptoms of COVID-19<sup>a</sup> | 0,353 (0,213-0,582) | 0,000 | 0,353 (0,211-0,588) | 0,000 |

<sup>a</sup> Yes: 1. No: 0. Abbreviations: RDT, rapid antigen detection test; CI, confidence interval, p-value, probability value; COVID-19, coronavirus disease 2019.

### RDT assessment and congruency with NAAT

Of all included participants, 400 (5.6%) subjects had an incongruent NAAT and RDT result, i.e. a positive NAAT result with a negative or invalid RDT result, or a negative NAAT result with a positive or invalid RDT result. Of those, 266 (66.5%) participants uploaded a photo of their RDT to the cloud. In 25 (9.4%) out of a total of 266 uploaded photos of incongruent results, the RDT was assessed differently by the reviewer than the HCW. Increasing HCW age was associated with reading errors among all incongruent test results (p 0,03).

### Questionnaire data

Additional data concerning the questionnaire of the included participants are shown in Table 7. Medians in all three study groups are similar for the perceived difficulty of the RDT. The least doubt about the RDT is observed in study group 3. Participants were most likely to use the test again when they had COVID-19 like symptoms or recommend the test to a colleague in study group 2 and study group 3.

**Table 7.**

|  | Study group |  |  | Overall |
| --- | --- | --- | --- | --- |
|  | Group 1* | Group 2** | Group 3*** |  |
| Test difficulty level, median (IQR) | 8 (6-8) | 8 (7-9) | 8 (6-8) | 8 (6-8) |
| Doubt about RDT result, n (%) |  |  |  |  |
| No doubt | 1909 (66,0) | 1092 (69,7) | 1453 (73,6) | 4454 (69,2) |
| Some doubt | 939 (32,5) | 449 (28,7) | 500 (25,3) | 1888 (29,3) |
| Serious doubt | 45 (1,6) | 25 (1,6) | 21 (1,1) | 91 (1,4) |
| Self-test again when COVID-19-like symptoms, n (%) |  |  |  |  |
| Disagree | 231 (7,3) | 80 (4,7) | 80 (3,7) | 391 (5,6) |
| Partly disagree | 133 (4,2) | 50 (2,9) | 84 (3,9) | 267 (3,8) |
| Partly agree | 583 (18,5) | 309 (18,2) | 369 (17,1) | 1261 (18,0) |
| Agree | 2202 (69,9) | 1258 (74,1) | 1622 (75,3) | 5082 (72,6) |
| Recommend test to colleague, n (%) |  |  |  |  |
| Disagree | 196 (6,2) | 67 (4,0) | 63 (2,9) | 326 (4,7) |
| Partly disagree | 125 (4,0) | 47 (2,8) | 85 (4,0) | 257 (3,7) |
| Partly agree | 636 (20,2) | 357 (21,1) | 407 (18,9) | 1400 (20,0) |
| Agree | 2191 (69,6) | 1222 (72,2) | 1596 (74,2) | 5009 (71,6) |

\*BD-RDT with combined oropharyngeal - mid-turbinate nasal sampling. \*\* BD-RDT with mid-turbinate nasal sampling only. \*\*\*Roche-RDT with combined oropharyngeal - mid-turbinate nasal sampling. Abbreviations: IQR, interquartile range; RDT, rapid antigen detection test; COVID-19, coronavirus disease 2019.

## Discussion

SARS-CoV-2 rapid antigen tests performed by HCW as self-tests were able to detect most infectious COVID-19 cases. The results were better when combined oropharyngeal - mid-turbinate samples were used compared to mid-turbinate samples only. Also, the sensitivity varied between different brands of RDT. This study shows that RDT’s can be used as self-tests but that there are certain limitations considering the sensitivity.

In general, RDT’s perform well in individuals with a high SARS-CoV-2 viral load which is usually present in the pre-symptomatic and early symptomatic phase – the first 5-7 days – of COVID-19 (11,12). Multiple validation studies for RDT have been published in which a higher sensitivity is observed at low Ct-values (high viral loads) for different RDT brands and different qRT-PCR platforms (1-3, 13-17). It is a point of discussion above which viral load and therefore below which Ct-value people are likely to be infectious. After infection with SARS-CoV-2, viral RNA can be detected by qRT-PCR during a long time, even if the infected person is no longer infectious (18, 19). Taking the viral load of SARS-CoV-2 as a proxy for infectiousness – by determining the viral load cut-off above which 95% of qRT-PCR positive samples had a positive virus culture – and extrapolating it to a cut-off Ct-value for comparable PCR platforms is the most feasible and closest to the true rate of infectiousness (6, 10, 20).

When adjusted for infectiousness for all qRT-PCR results of samples processed in the cobas^®^ 6800/8800 the sensitivity clearly increased for each group. The sensitivities were 82,2%, 61,9% and 90,2% for study group 1, study group 2 and study group 3, respectively. The increased sensitivity after adjustment for infectiousness is in line with previous studies (6, 10). Specificity decreased slightly in all study groups after adjustment for infectiousness. This is a consequence of a number of previously true-positive RDT results compared with qRT-PCR results with viral loads below the cut-off that now pertained to the false-positive group. As WHO stated, RDTs would need a sensitivity ≥80% and a specificity ≥97% (21). Both BD-RDT and Roche-RDT with combined oropharyngeal - mid-turbinate sampling meet this requirement and BD-RDT in the case of mid-turbinate sampling only does not meet this WHO recommendation on sensitivity. Addition of an oropharyngeal swab to a mid-turbinate nasal sample clearly improves the yield.

We found that reliability of self-testing with RDT among HCW seems to depend on the type of RDT, sampling method of the mucosal specimen and the presence of COVID-19 like symptoms at the time of testing. Participants that assessed a Roche-RDT were significantly less likely to have a false-negative RDT result compared to participants that assessed a BD-RDT (p <0.01). Subjects that performed combined oropharyngeal – mid-turbinate nasal sampling were significantly less likely to have a false-negative RDT result compared to participants that performed mid-turbinate nasal sampling only (p <0,05). By contrast, it appears from the literature that nasal mid-turbinate self-sampling versus nasopharyngeal sampling performed by a trained subject would not have an effect on sensitivity of an RDT (7, 22). To our knowledge, this is the first clinical study that has shown a significant difference in the reliability of the RDT type and sampling method when applied as self-test. Also, the presence of COVID-19 like symptoms at the time of testing appeared to be a factor in the multivariable regression analysis that reduces the probability of a false-negative RDT (p <0,01). This is consistent with the finding of Thirion-Romero et al. that the presence of symptoms was a predictor of positivity for the Panbio RDT (23).

Our study was conducted in the Netherlands in a time frame just before a transition from the original Wuhan strain to the Alpha variant (9). Recent data of the FDA suggested that RDT do detect the omicron variant but may have reduced sensitivity (24). In particular, the media raised the question whether this alleged reduced sensitivity of the RDT in the SARS-CoV-2 Omicron variant might be due to the sampling method. Omicron is said to be more detectable by an RDT with a combined oropharyngeal - nasal sampling than a nasal sampling only. Our study shows that a significant difference in RDT sensitivity is demonstrable with a difference in the sampling method. Whether the size of this difference is applicable with the emergence of the omicron variant is still to be determined.

Self-test RDT accuracy among HCW is not affected by reading errors, nor does it seem to matter for the performance of the test whether someone is medically trained or not. In our study hardly any self-reading errors were made. The total percentage of reading errors of all inclusions is estimated to be 0.5%, assuming that all congruent results have been read correctly and that the percentage of reading error in the group of incongruent results with no photo uploaded is also 9.5%. Little has been described in literature about self-reading of antigen tests in general. It can be assumed that HCW may have more experience with the interpretation of these types of tests than e.g. laypersons. Lindner et al. reported that the inter-rater reliability in a double reading of an RDT between laypersons and professionals was very high (kappa 0,98) (8) and Cassuto et al stated that interpretation errors in RDT also occur very rarely in laymen (25).

Our study had a number of limitations. First was the number of different NAAT platforms used. Except for the cobas^®^ 6800/8800 platform the correlation between Ct-value produced by qRT-PCR platforms and the SARS-CoV-2 viral load is not known. Further, not every NAAT platform renders a Ct-value as a result and for that we were unable to extrapolate those NAAT results to viral loads. As a result, we had to omit a large amount of subjects from our logistic regression analysis. We could only take the data that was run on the cobas^®^ 6800/8800 platform, because for this platform we previously determined the correlation between viral load and Ct-value (10). In future research, it would be advisable - in the case of a multicenter study - to have a known and described cut-off point for each platform used or to run all samples primarily on a single platform to facilitate this comparison process. The most accurate would be to perform virus cultures, although this will not always be feasible. Secondly, different methods of upper respiratory specimen collection, as nasopharyngeal or mid-turbinate either with or without oropharyngeal sampling for NAAT were included when performed by trained personnel, according to the local protocol in de participating center which makes the comparison of results of the reference test between the centers not completely reliable. Thirdly, the experience with the test procedure may grow over time leading to a better performance for tests performed at the end of the study. As the procedures were not performed in the same period this may play a role. However, this may be of limited importance as the results of the BD-RDT in period two were worse than in period one.

In conclusion, SARS-CoV-2 rapid antigen tests have proven to be able to detect infectious individuals when upper respiratory specimen is collected with a combined oropharyngeal - mid-turbinate sampling method. Reliability of self-testing with RDT among HCW seems to depend on the type of RDT, sampling method and the presence of COVID-19 like symptoms at the time of testing. Self-test RDT accuracy among HCW is not affected substantially by reading errors, nor whether or not a HCW is medically trained. This makes self-testing using RDT a useful tool to detect infectious individuals.

## Data Availability

All data produced in the present study are available upon reasonable request to the authors.

## Acknowledgement

We would like to thank Ons Tweede Thuis for participating in the study. I would also like to thank Berit Hol for the effort put into the instruction forms.

## References

1. Van der Moeren N, Zwart VF, Lodder EB, Van den Bijllaardt W, Van Esch Hrjm, Stohr Jjjm, et al. Evaluation of the test accuracy of a SARS-CoV-2 rapid antigen test in symptomatic community dwelling individuals in the Netherlands. PLOS ONE. 2021;16(5):e0250886.

2. Igloi Z, Velzing J, van Beek J, van de Vijver D, Aron G, Ensing R, et al. Clinical Evaluation of Roche SD Biosensor Rapid Antigen Test for SARS-CoV-2 in Municipal Health Service Testing Site, the Netherlands. Emerging Infectious Disease journal. 2021;27(5):1323.

3. Gremmels H, Winkel BMF, Schuurman R, Rosingh A, Rigter NAM, Rodriguez O, et al. Real-life validation of the Panbio™ COVID-19 antigen rapid test (Abbott) in community-dwelling subjects with symptoms of potential SARS-CoV-2 infection. EClinicalMedicine. 2021;31:100677.

4. Venekamp RP, Veldhuijzen IK, Moons KGM, van den Bijllaardt W, Pas SD, Lodder EB, et al. Diagnostic accuracy of three prevailing rapid antigen tests for detection of SARS-CoV-2 infection in the general population: cross sectional study. medRxiv. 2021:2021.11.19.21266579.

5. Brümmer LE, Katzenschlager S, Gaeddert M, Erdmann C, Schmitz S, Bota M, et al. Accuracy of novel antigen rapid diagnostics for SARS-CoV-2: A living systematic review and meta-analysis. PLOS Medicine. 2021;18(8):e1003735.

6. Stohr Jjjm, Zwart VF, Goderski G, Meijer A, Nagel-Imming CRS, Kluytmans-van den Bergh MFQ, et al. Self-testing for the detection of SARS-CoV-2 infection with rapid antigen tests for people with suspected COVID-19 in the community. Clinical Microbiology and Infection. 2021.

7. Klein JAF, Krüger LJ, Tobian F, Gaeddert M, Lainati F, Schnitzler P, et al. Head-to-head performance comparison of self-collected nasal versus professional-collected nasopharyngeal swab for a WHO-listed SARS-CoV-2 antigen-detecting rapid diagnostic test. Med Microbiol Immunol. 2021;210(4):181–6.

8. Lindner AK, Nikolai O, Rohardt C, Kausch F, Wintel M, Gertler M, et al. Diagnostic accuracy and feasibility of patient self-testing with a SARS-CoV-2 antigen-detecting rapid test. J Clin Virol. 2021;141:104874.

9. Rijksoverheid Coronadashboard. Besmettingen. Varianten van het coronavirus door de tijd heen. https://coronadashboard.rijksoverheid.nl/landelijk/varianten.

10. Schuit E, Veldhuijzen IK, Venekamp RP, van den Bijllaardt W, Pas SD, Lodder EB, et al. Diagnostic accuracy of rapid antigen tests in asymptomatic and presymptomatic close contacts of individuals with confirmed SARS-CoV-2 infection: cross sectional study. BMJ. 2021;374:n1676.

11. Weiss A, Jellingsø M, Sommer MOA. Spatial and temporal dynamics of SARS-CoV-2 in COVID-19 patients: A systematic review and meta-analysis. EBioMedicine. 2020;58:102916-.

12. Arons MM, Hatfield KM, Reddy SC, Kimball A, James A, Jacobs JR, et al. Presymptomatic SARS-CoV-2 Infections and Transmission in a Skilled Nursing Facility. New England Journal of Medicine. 2020;382(22):2081–90.

13. Albert E, Torres I, Bueno F, Huntley D, Molla E, Fernández-Fuentes MÁ, et al. Field evaluation of a rapid antigen test (Panbio™ COVID-19 Ag Rapid Test Device) for COVID-19 diagnosis in primary healthcare centres. Clin Microbiol Infect. 2021;27(3):472.e7-.e10.

14. Krüttgen A, Cornelissen CG, Dreher M, Hornef MW, Imöhl M, Kleines M. Comparison of the SARS-CoV-2 Rapid antigen test to the real star Sars-CoV-2 RT PCR kit. J Virol Methods. 2021;288:114024-.

15. Krüger LJ, Gaeddert M, Tobian F, Lainati F, Gottschalk C, Klein JAF, et al. The Abbott PanBio WHO emergency use listed, rapid, antigen-detecting point-of-care diagnostic test for SARS-CoV-2— Evaluation of the accuracy and ease-of-use. PLOS ONE. 2021;16(5):e0247918.

16. Ngo Nsoga MT, Kronig I, Perez Rodriguez FJ, Sattonnet-Roche P, Da Silva D, Helbling J, et al. Diagnostic accuracy of Panbio rapid antigen tests on oropharyngeal swabs for detection of SARS-CoV-2. PLOS ONE. 2021;16(6):e0253321.

17. Kweon OJ, Lim YK, Kim HR, Choi Y, Kim M-C, Choi S-H, et al. Evaluation of rapid SARS-CoV-2 antigen tests, AFIAS COVID-19 Ag and ichroma COVID-19 Ag, with serial nasopharyngeal specimens from COVID-19 patients. PLOS ONE. 2021;16(4):e0249972.

18. van Kampen JJA, van de Vijver DAMC, Fraaij PLA, Haagmans BL, Lamers MM, Okba N, et al. Duration and key determinants of infectious virus shedding in hospitalized patients with coronavirus disease-2019 (COVID-19). Nature Communications. 2021;12(1):267.

19. Crozier A, Rajan S, Buchan I, McKee M. Put to the test: use of rapid testing technologies for covid-19. BMJ. 2021;372:n208.

20. Guglielmi G. Rapid coronavirus tests: a guide for the perplexed. Nature. 09 February 2021(590):202–5.

21. World Health Organization. WHO Interim Guidance 11 September 2020. Antigen-detection in the Diagnosis of SARS-CoV-2 Infection using Rapid Immunoassays. https://apps.who.int/iris/bitstream/handle/10665/334253/WHO-2019-nCoV-Antigen_Detection-2020.1-eng.pdf?sequence=1&isAllowed=y

22. Nikolai O, Rohardt C, Tobian F, Junge A, Corman VM, Jones TC, et al. Anterior nasal versus nasal mid-turbinate sampling for a SARS-CoV-2 antigen-detecting rapid test: does localisation or professional collection matter? Infect Dis (Lond). 2021;53(12):947–52.

23. Thirion-Romero I, Guerrero-Zúñiga DS, Arias-Mendoza DA, Cornejo-Juárez DDP, Meza-Meneses DP, Torres-Erazo DDS, et al. Evaluation of Panbio rapid antigen test for SARS-CoV-2 in symptomatic patients and their contacts: a multicenter study. Int J Infect Dis. 2021;113:218–24.

24. U.S. Food&Drug Administration. SARS-CoV-2 Viral Mutations: Impact on COVID-19 Tests. 28 December 2021. https://www.fda.gov/medical-devices/coronavirus-covid-19-and-medical-devices/sars-cov-2-viral-mutations-impact-covid-19-tests.

25. Cassuto NG, Gravier A, Colin M, Theillay A, Pires-Roteira D, Pallay S, et al. Evaluation of a SARS-CoV-2 antigen-detecting rapid diagnostic test as a self-test: Diagnostic performance and usability. Journal of Medical Virology. 2021;93(12):6686–92.

