## Supplementary Methods 1 for "Performance of Various Lateral Flow SARS-CoV-2 Antigen Self Testing Methods in Healthcare Workers: a Multicenter Study"

Note: The original document below is in Dutch. This is a version translated into English.

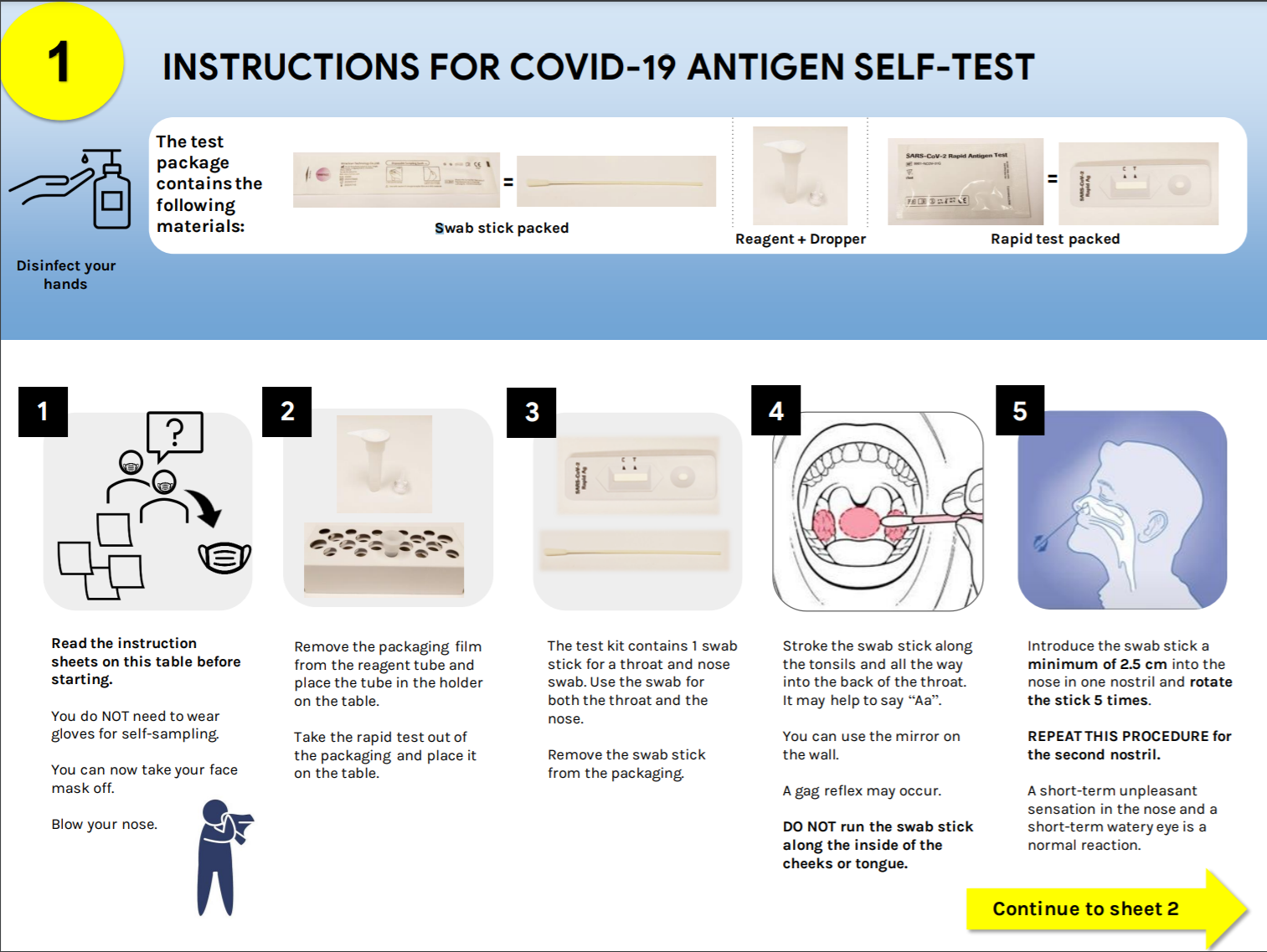

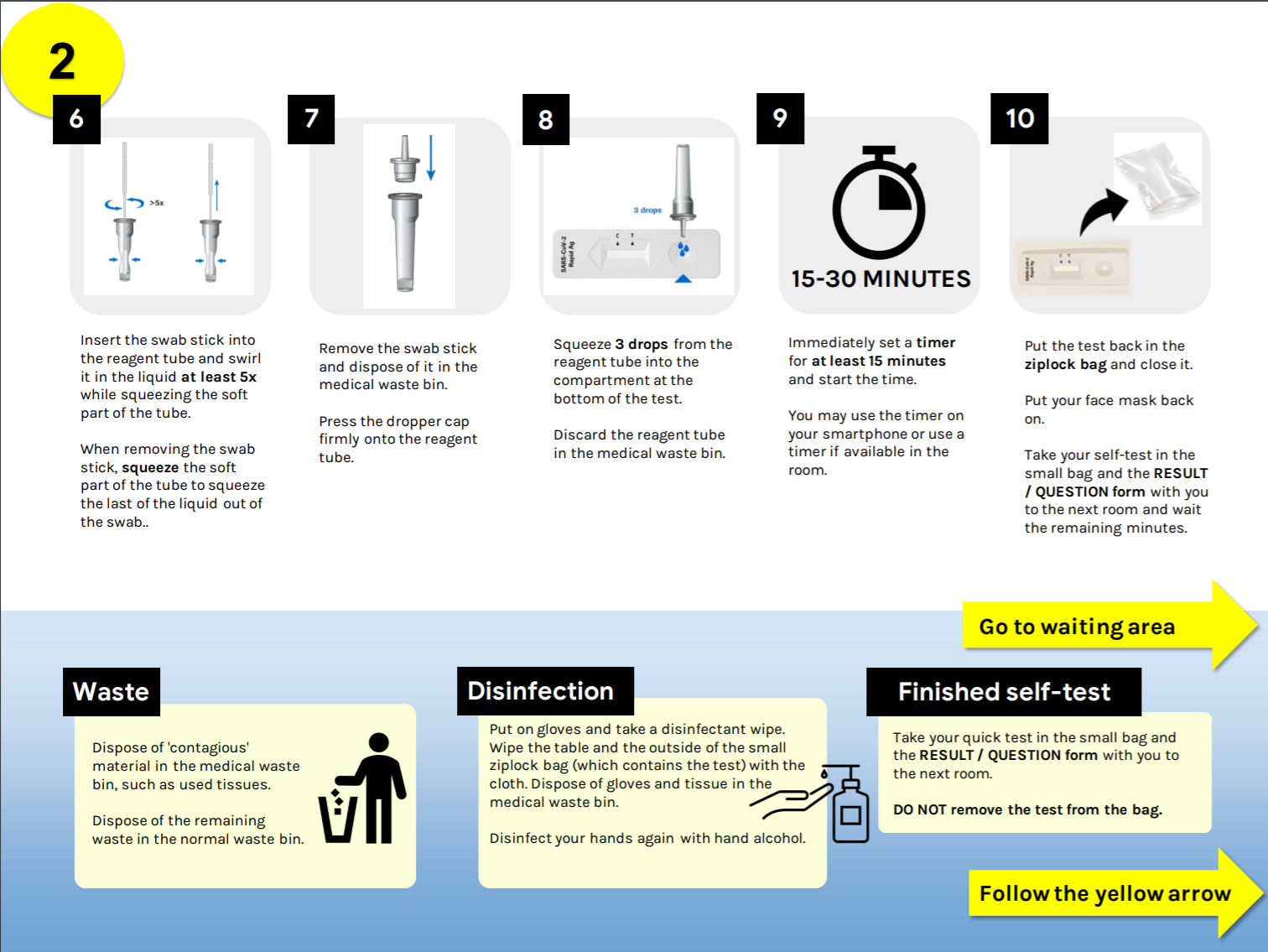

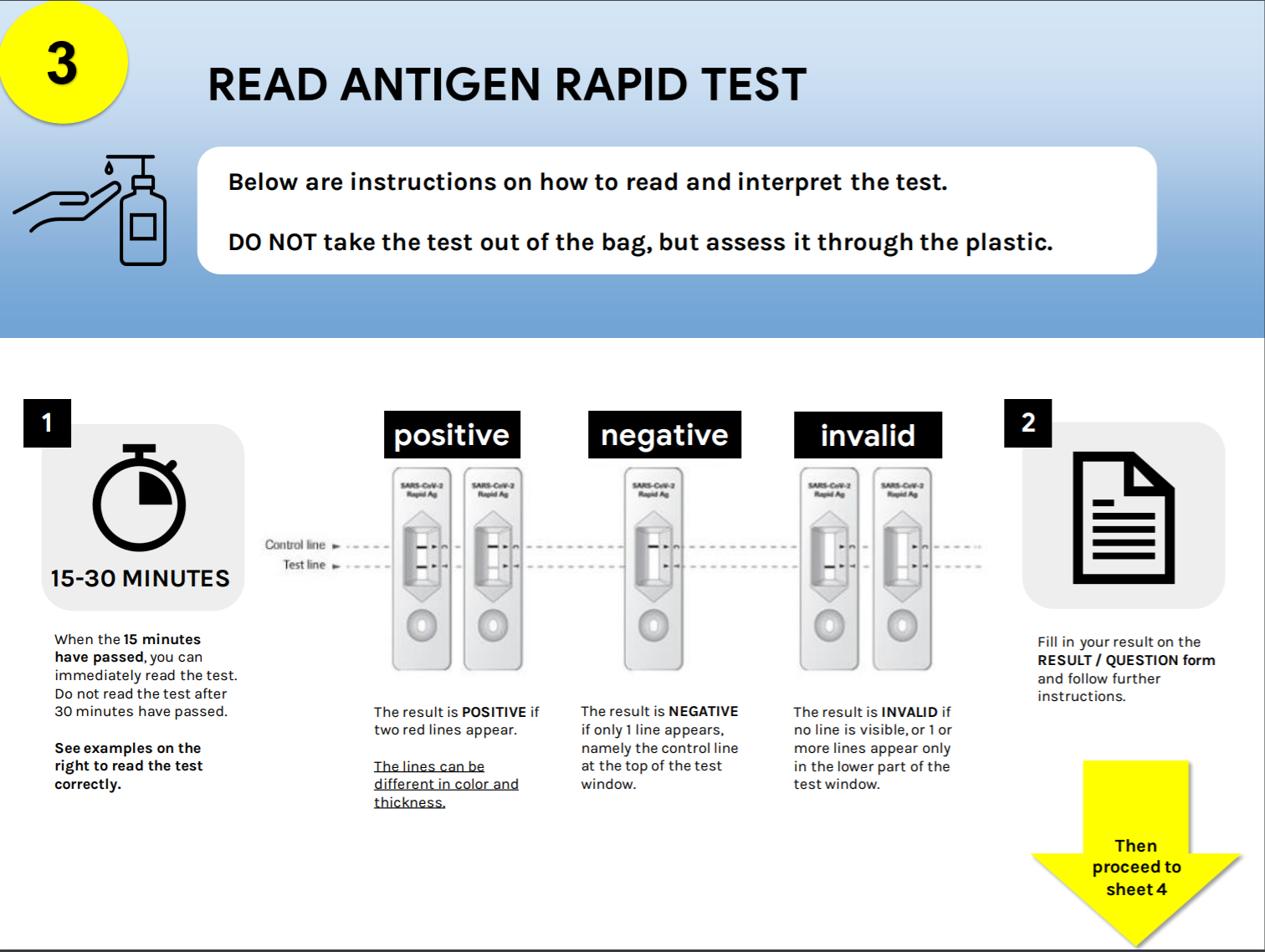

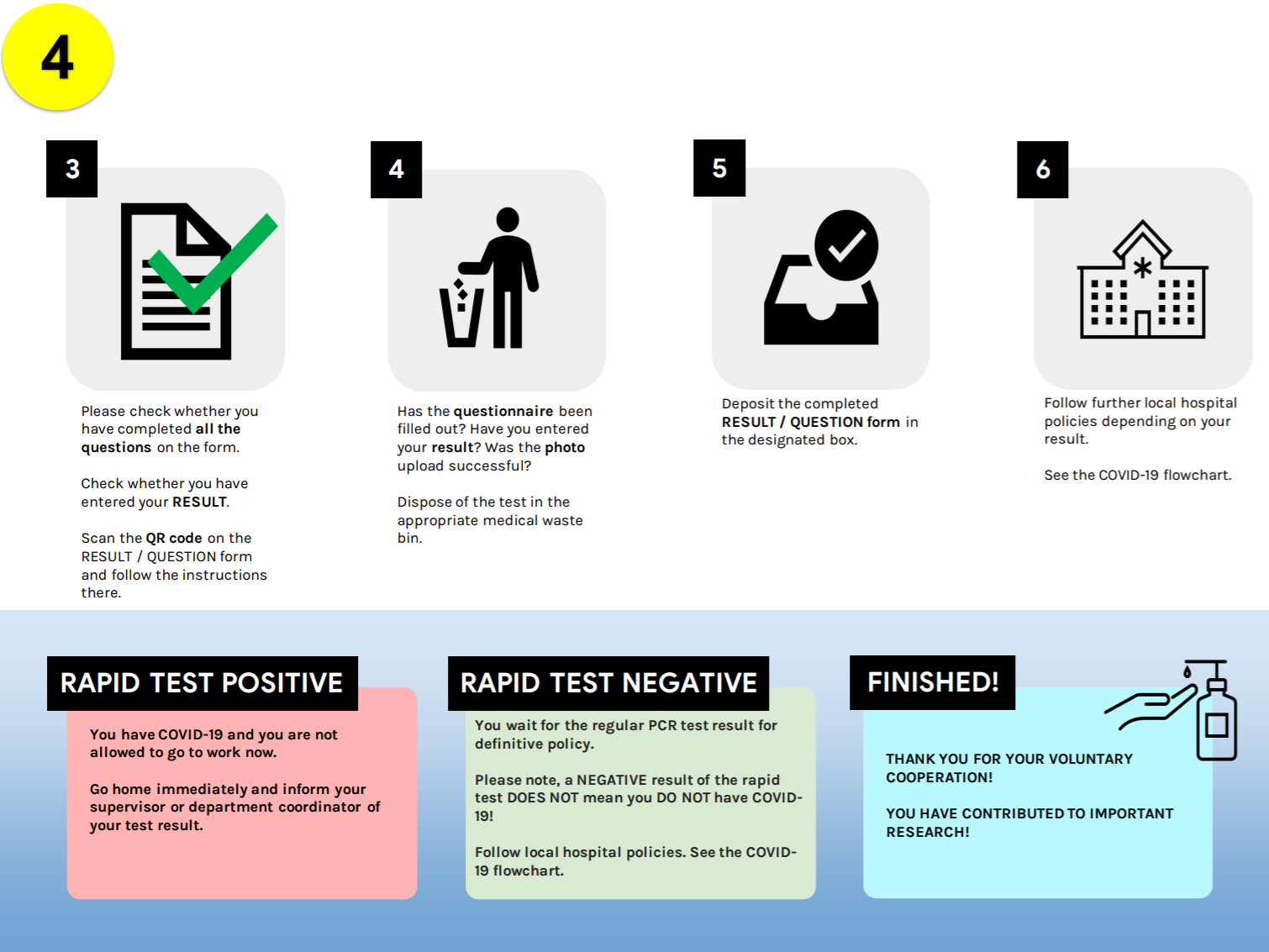
