## Supplementary Methods 2 for "Performance of Various Lateral Flow SARS-CoV-2 Antigen Self Testing Methods in Healthcare Workers: a Multicenter Study"

Note: The original document below is in Dutch. This is a version translated into English.

1. What is your age?

2. What is your gender?

3. What is your group of profession? (Tick one answer)

- Medical (including nurse, doctor, laboratory, outpatient assistants)
- Supportive (including facilities, housekeeping, transport, food)
- Office function (including secretarial, administrative, human resource)
- Volunteer (including host/hostess)

4. In the past 3 weeks, have you had any symptoms related to COVID-19*?

- Yes, please enter the date on which these symptoms started.
- No

* Cold, sore throat, cough, sneezing, shortness of breath, fever, loss of smell or taste, general malaise, headache, muscle pain or inexplicable diarrhoea.

5. Do you currently (still) have symptoms?

- Yes
- No

6. On a scale of 1 to 10 (with 1 being the most difficult and 10 being the easiest), how difficult or easy do you find performing the test.

7. What is the result of your self-test?

- Positive
- Negative
- Invalid

8. Do you doubt your self-assessed result?

- I have no doubt
- I have some doubt
- I have serious doubt

9. Do you disagree or agree with the following two statements:

I would like to do this self-test again if I had COVID-19 symptoms.

- Disagree
- Partly disagree
- Partly agree
- Agree

I would recommend this self-test to a colleague.

- Disagree
- Partly disagree
- Partly agree
- Agree
