## Supplementary Methods 3 for "Performance of Various Lateral Flow SARS-CoV-2 Antigen Self Testing Methods in Healthcare Workers: a Multicenter Study"

Viral load calculation

Viral loads for samples processed on the cobas^®^ 6800/8800 platform were calculated according to Schuit et al. (1). For that study a standard curve was created by the Erasmus MC viroscience laboratory, by testing dilutions of a specific quantified E-gene transcript (primary standard) as made available by the European Virus Archive (2). The qRT-PCR used was based on the publication of Coreman et al. (3). The mathematical formula describing the relation between Ct-value and number of E-gene copies was then used to quantify the viral load in a secondary standard that was prepared out of cell-cultured SARS-CoV-2 virus. A new standard curve was then prepared by testing of a serial dilution of the secondary standard on the cobas^®^ 6800 platform.

By testing the same SARS-CoV-2 panel obtained from the National Public Health Institute at Microvida laboratory and Erasmus MC it was determined that both cobas^®^ 6800 machines resulted comparable Ct-values for the same samples. For the current study, Microvida laboratory was the only laboratory using the cobas^®^ platform. Validation of the cobas^®^ 8800 machine that was installed during the study period determined that also this machine resulted comparable Ct-values when the same sample is tested.

To account for a variable amount of medium/reagent volume per swab this factor is included in the Ct-value – viral load (copies/ml) conversion formula. This formula was 62.5*𝑒^(43.1-Ct)/1.607^/3⅓ for an initial sample volume of 1.8 ml and 62.5*𝑒^(43.1-Ct)/1.607^ for an initial sample volume of 6 ml.

**References**

(1) Schuit E, Veldhuijzen I K, Venekamp R P, van den Bijllaardt W, Pas S D, Lodder E B et al. Diagnostic accuracy of rapid antigen tests in asymptomatic and presymptomatic close contacts of individuals with confirmed SARS-CoV-2 infection: cross sectional study BMJ 2021; 374 :n1676 doi:10.1136/bmj.n1676.

(2) Archive EV. Wuhan coronavirus 2019 E gene control 2020 [Available from: https://www.european-virus-archive.com/nucleic-acid/wuhan-coronavirus-2019-e-gene-control accessed June 8 2021 2021.

(3) Corman VM, Landt O, Kaiser M, et al. Detection of 2019 novel coronavirus (2019-nCoV) by real-time RT-PCR. Euro Surveill 2020;25.
